# Effects of exercise on sleep in children with overweight/obesity: A randomized clinical trial

**DOI:** 10.1101/2022.09.23.22280266

**Authors:** Lucia V. Torres-Lopez, Jairo H. Migueles, Cristina Cadenas-Sanchez, Marcus Bendtsen, Pontus Henriksson, Jose Mora-Gonzalez, Marie Löf, Jean-Philippe Chaput, Francisco B. Ortega

## Abstract

**Objectives:** To examine 1) the chronic effects of a 20-week physical exercise program on device-assessed sleep and on sleep-disordered breathing (SDB); and 2) whether attending to a session of the exercise program had effects on device-assessed sleep the subsequent night in children with overweight/obesity.

**Methods:** A total of 99 children with overweight/obesity (n=47 in the exercise group) participated in this secondary analysis of the ActiveBrains randomized clinical trial. The exercise program included a combination of aerobic and resistance exercise training, 3-5 days/week (90 min/session). The control group was asked to maintain usual lifestyle. Sleep outcomes were measured using wrist actigraphy and included: total sleep time, total time in bed, sleep efficiency, and wake after sleep onset (WASO) time. SDB was assessed via the Paediatric Sleep Questionnaire.

**Results:** The ActiveBrains exercise program had a statistically significant effect on WASO time (−10.8 min/day, -0.5 standard deviations (SD), *P*=0.040). Furthermore, we observed a small non-statistically significant effect on sleep efficiency (+2%, 0.4 SD, *P*=0.1). No other chronic effects were observed on the other sleep outcomes. The nights (at maximum of 4) after attending the ActiveBrains exercise sessions, children showed higher total sleep time (+8 min, *P*=0.17), sleep efficiency (+1%, *P*=0.15), and lower WASO time (−6 min, *P*=0.18) although did not reach statistical significance.

**Conclusion:** A 20-week physical exercise program reduced WASO time in children with overweight/obesity in comparison with control group peers, yet we did not observe effects on SDB. Future randomized trials that include a clinical sample of children with poor sleep health at baseline are needed to better appreciate the role of exercise in sleep health.

**Clinal Trial Registration:** The ActiveBrains project is registered at ClinicalTrials.gov, no. NCT02295072. Data Sharing Statement: We did not obtain children’s parents consent to widely share the data nor was it included in the IRB protocol.

**What’s known on this subject:** Sleep is an essential component in children’s health. Previous researchers have found positive effects of exercise on sleep in adolescents with obesity, but the literature in childhood obesity is lacking.

**What this study adds:** A physical exercise program showed positive effects on device-assessed sleep habits in children with overweight/obesity, yet not on sleep-disordered breathing. Exercise programs should be promoted in children to improve their sleep health, especially in those with overweight/obesity.

## Introduction

Sleep is essential for children’s health and well-being.^1^ The current lifestyles (e.g., late-night screen time, energy drinks, and delayed bedtimes) are compromising the sleep habits in children.^2^ As a result, the attainment to the sleep recommendations (uninterrupted 9-11 h of sleep/night) is decreasing in children.^3^ The estimated prevalence of 6-to-13 year-old children sleeping less than 9-11h/day is 98.1%, as measured with accelerometers, and 18.1% as reported in diaries;^4^ and the prevalence of sleep-disordered breathing (SDB) is around 4-11% in children.^5^ Shorter sleep duration has been related to worse physical and mental health in children and adolescents^6^, and poorer sleep outcomes (i.e., sleep duration and sleep patterns) appear to be associated with childhood obesity.^7–9^ As an example, we have previously observed a link between device-assessed sleep outcomes^10^ and SDB^11^ with brain health (i.e., gray matter volume and academic performance) in children with overweight/obesity. Given the well-contrasted multi-organ benefits of physical exercise,^12^ it is of interest to investigate whether physical exercise improves sleep in children with overweight/obesity.

A large number of studies have supported the positive chronic and acute effects of physical exercise on device-assessed sleep in adults.^13^ However, the literature focused on the effects of physical exercise on sleep in the pediatric population is scarce. To date, only one randomized clinical trial (RCT) investigated the effect of exercise intervention on device-assessed sleep in children with autism spectrum disorders.^14^ In obesity, the available trials on exercise and sleep are limited to adolescents,^15^ while the studies in childhood are all observational and focused on lifestyle physical activity and sleep.^16,17^ In regards to SDB, few studies showed that engaging in at least 20/40 minutes per day of vigorous aerobic exercise appeared to be beneficial in palliating symptoms of SDB in children 7 to 11 year-old with overweight,^18^ but not in obese children with SDB.^19^ There is a need of RCTs on the effects of exercise on sleep in pediatric obesity.

Few studies examined the chronic and acute effects of exercise on sleep outcomes (i.e., sleep time and quality) in adolescents with obesity.^15,20^ This information would inform public health recommendations and practice. Therefore, the aims of this study were to examine 1) the chronic effects of a 20-week physical exercise program on device-assessed sleep, and on SDB; and 2) whether attending to a session of the exercise program had effects on device-assessed sleep the subsequent night in children with overweight/obesity.

## Methods

### Study design and participants

The ActiveBrains project was a RCT (Clinical Trial registration no. NCT02295072) which primary aim was to investigate the effect of a 20-week exercise program on brain health indicators in children with overweight/obesity.^21,22^ This study examined the ActiveBrains RCT effects on sleep (secondary outcomes). Details on the design, eligibility, characteristics, and effects on the primary outcomes can be found elsewhere.^21,22^ In brief, 109 children (8-11 years) with overweight/obesity took part in this RCT.^21,22^ After baseline data collection, participants were randomly allocated to either the wait-list control group (N=52) or exercise program (N=57) using a computer-based simple randomization procedure in a ratio of 1:1 by a person not involved in the assessments. Participants in the wait-list control group continued with their usual routines. **Figure 1** depicts the participants flowchart. Data were collected from November 2014 to June 2016 in Granada (Spain). The ActiveBrains RCT^21,22^ was conducted according to the Declaration of Helsinki and the study protocol was approved by the Ethics Committee on Human Research (CEIH) of the University of Granada.

**Figure 1.**
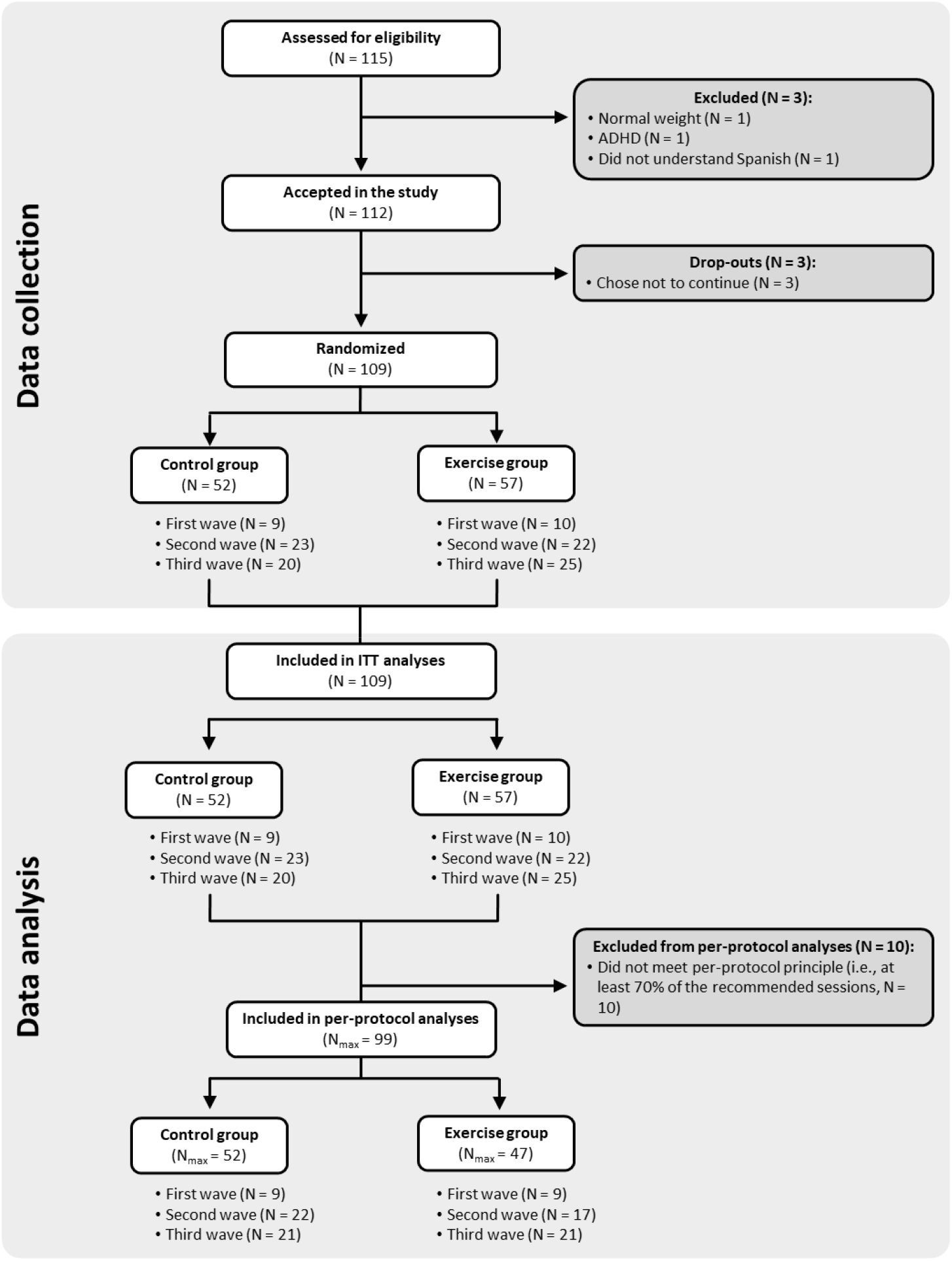
Flowchart of the study participants. ADHD: Attention-deficit hyperactivity disorder; ITT: Intention-to-treat. For final ITT analyses, those participants that left the study during the physical exercise program or did not complete the post-exercise program assessments were imputed (see Statistical section). N_max_: Maximum N for analyses, it changes depending on the variable.

Written informed consent was obtained by the legal guardians or parents of all participants, who were informed of the purpose of the study and provided informed assent. Consolidated Standards of Reporting Trials (CONSORT) guidelines were followed for the reporting of trial results.^23^

### Physical exercise program

The ActiveBrains physical exercise program had a duration of 20 weeks and was based on the international guidelines on physical activity for children.^24^ Participants were recommended to attend a minimum of 3 out of 5 sessions/week. The exercise program was based on aerobic and resistance exercise. Each session lasted 90 min (i.e., 5-min warm-up; 60-min aerobic exercise; 20-min resistance exercise; and 5-min cool-down).

### Measurements

Both control and exercise groups were assessed at baseline and post-intervention. Sleep and physical activity were additionally monitored in the middle of the exercise program (week 10^th^) in both groups. Assessors were not fully blinded to the participant’s group allocation due to practical reasons. More information about the measurements can be found elsewhere.^21^

### Sleep-disordered breathing

The children’s legal guardians completed the Spanish version of the Paediatric Sleep Questionnaire (PSQ) to evaluate SDB, which has shown high reliability and internal consistency^25^ as well as validity for the identification of SDB.^26,27^ The sleep-related breathing disorders (SRBD) scale (range: 0 to 1) was used to quantify SDB, with higher scores indicating greater breathing severity.^27^ This scale consists of 22 closed response items of the reduced version of the PSQ, which were divided into three subscales:^26,27^ snoring, sleepiness and inattention/hyperactivity. Further details on the questionnaire can be found elsewhere.^11,28^

### Device-assessed sleep

Sleep was monitored with non-dominant wrist-worn accelerometers (ActiGraph GT3X+, Pensacola, FL, USA) for 7 consecutive days (24h/day). Participants reported information on the time they went to bed and woke up every day. Accelerometers were initialized to record accelerations at 100 Hz, and the files were processed in the R package GGIR (v.1.5.12, https://www.cran.r-project.org/).^29^ More detailed information on accelerometer data processing for device-assessed sleep outcomes analysis can be found elsewhere.^30^ The algorithm developed by Sadeh et al.^31^ was applied within the total time in bed frame to classify every minute as “sleep” or “wake” time. Device-assessed sleep outcomes included indicators of sleep timing, sleep duration (i.e., total sleep time and total time in bed), and sleep quality (i.e., sleep efficiency and wake after sleep onset [WASO] time). A valid day was considered if the participant registered at least 4 days (≥ 16 hours/day), including a minimum of 1 weekend day.

### Statistical analysis

Descriptive characteristics of the study sample are shown as mean and standard deviations (SD) or percentages and frequencies.

#### Chronic effect of the physical exercise program on sleep

The chronic effects of the ActiveBrains exercise program on each sleep-related outcome were conducted according to the per-protocol principle (i.e., attending at least 70% of the sessions) using an analysis of covariance (ANCOVA). Per-protocol analyses were the main analyses in this trial to investigate the efficacy rather than effectiveness of our intervention (i.e., effects of the preconceived intervention). Post-exercise sleep-related outcome values were included as dependent variables, group (i.e., exercise vs. control) as fixed factor, and baseline outcome data as covariates.^32^ Raw and standardized (z-scores) estimates are provided, with the post-exercise z-scores calculated relative to the baseline mean and SD as an indicative of the change.^21,32^ A difference of 0.2 SDs was considered a small, 0.5 SDs medium, and 0.8 SDs large effect sizes.^33^ Differences exceeding 0.2 Cohen’s D were interpreted as minimum meaningful effect.^34^

Exploratory analyses were conducted following the intention-to-treat-principle, including all the recruited participants, and imputing the missing values using predictive mean matching multiple imputations.^35^ We checked that missing data was missing at random prior to performing the multiple imputation.

#### Acute effect of the physical exercise program on sleep

The acute effects of the ActiveBrains exercise program were investigated over the sleep outcomes from the subsequent nights (at maximum of 4 nights) to the sessions the participants attended versus the sleep outcomes from the nights after the days in which they did not attend to the sessions (at week 10^th^, in the middle of the intervention). This analysis only included participants allocated to the exercise group. We excluded Fridays, Saturdays and Sundays from the dataset given the expected changes in the sleep patterns during weekends.^36^ Linear regression mixed models were used with the attendance indicator (0=non-attendance to exercise program, 1=attendance) as fixed factor, the ID as random factor, and sleep indicators as outcomes. Random and fixed intercepts were defined to investigate the within-group (attendance and non-attendance) and within-participant effect of the exercise session, respectively.

Analyses were performed using the Statistical Package for Social Science (IBM SPSS Statistics for Windows, version 22.0, Armonk, NY) and R v.3.4.1 (https://cran.r-project.org/). The level of significance was set at P < 0.05.

To better understand the acute effects of the exercise program on sleep, we investigated whether the children compensated the amount of exercise performed in the ActiveBrains session with activities that they usually perform in their daily life. For such purpose, we performed a 1-dimension curve analysis using statistical parametric mapping software (SPM1D; Wellcome Department of Cognitive Neurology, London, UK) to verify whether acceleration values identified a significant physical activity increment in the days attending to the physical exercise sessions compared to the days not attending. From Monday to Thursday, mean acceleration curves were presented from midnight to the following midnight (i.e., 24h curves). T-tests were used to determine significant differences between the curves for attendance and non-attendance exercise group throughout the day.

## Results

### Descriptive characteristics

A total of 99 children with overweight/obesity (n=47 in the exercise group) were included in the per-protocol analysis (90% of the participants initially enrolled); the children who did not adhere to the protocol were not different in baseline characteristics from the included children (**Figure 1**). Descriptive baseline characteristics are shown in **Table 1**.

**Table 1.** Descriptive baseline characteristics of the ActiveBrains participants meeting the per-protocol criteria.

|  | All |  | Exercise group |  | Control group |  |
| --- | --- | --- | --- | --- | --- | --- |
|  | N | Mean (SD) or % | N | Mean (SD) or % | N | Mean (SD) or % |
| Sex |  |  |  |  |  |  |
| Girls (n %) | 41 | 41.4 | 16 | 34 | 25 | 48.1 |
| Boys (n %) | 58 | 58.6 | 31 | 66 | 27 | 51.9 |
| Age (years) | 99 | 10.1 (1.1) | 47 | 10.0 (1.1) | 52 | 10.1 (1.1) |
| Weight (kg) | 99 | 56.5 (11.0) | 47 | 57.4 (12.7) | 52 | 55.7 (9.4) |
| Height (cm) | 99 | 144.6 (8.3) | 47 | 143.9 (8.9) | 52 | 145.2 (7.9) |
| *Parent education level (n%)* |  |  |  |  |  |  |
| Primary school | 65 | 65.7 | 33 | 70.2 | 32 | 61.5 |
| Secondary school | 16 | 16.2 | 6 | 12.8 | 10 | 19.2 |
| University degree | 18 | 18.2 | 8 | 17.0 | 10 | 19.2 |
| Peak height velocity (years) | 99 | -2.2 (1.0) | 47 | -2.4 (0.9) | 52 | -2.1 (1.1) |
| *Sleep-related breathing disorders (SRBD)* |  |  |  |  |  |  |
| SRBD scale (0 to 1) | 99 | 0.19 (0.13) | 47 | 0.19 (0.13) | 52 | 0.19 (0.14) |
| SRBD presence (%) |  | 16.2 |  | 12.8 |  | 19.2 |
| *Device-assessed sleep* |  |  |  |  |  |  |
| Total sleep time (min/day) | 97 | 462.5 (36.0) | 47 | 462 (33.3) | 50 | 463 (38.7) |
| Total time in bed (min/day) | 97 | 529 (31.0) | 47 | 524.6 (35.1) | 50 | 533.1 (26.4) |
| Sleep efficiency (%) | 97 | 85 (4.6) | 47 | 85.3 (4.0) | 50 | 84.8 (5.2) |
| WASO time (min/day) | 97 | 74.9 (22.0) | 47 | 73.9 (19.1) | 50 | 75.8 (24.5) |

Data analyses were primarily conducted under the per-protocol principle, i.e., attending to 70% of the sessions or keep the usual lifestyle for exercise and control groups, respectively. SRBD was measured using the Spanish version of the Paediatric Sleep Questionnaire. Device-assessed sleep was measured using wrist-worn accelerometer. WASO: wake after sleep onset

### Chronic effect of the physical exercise program on sleep

The effects of the ActiveBrains physical exercise program on sleep outcomes are presented in **Table 2**. The largest effect of the ActiveBrains physical exercise program was found on WASO time, with an intervention vs. control difference of -10.79 min/day (95% CI:-21.09 to -0.51; P=0.040), showing a difference of medium effect size in the exercise group compared to the control group (SD=-0.507 [95% CI:-0.989 to -0.024]). Furthermore, a smaller and non-significant positive effect was observed for sleep efficiency (+2% [95% CI:-0.330 to 3.671]; SD=0.398). We found no evidence of exercise effects either on the remaining device-assessed sleep outcomes (i.e., total sleep time, sleep efficiency, and total time in bed) nor on SDB (all P > 0.05). The intention-to-treat analyses showed similar results, yet of smaller magnitude, than per-protocol analyses (**Supplemental Table 1**). More participants in the exercise group showed meaningful changes (i.e., reduction of ≥ 0.2 SDs) compared to the control group in WASO time (31% vs. 19%, P=0.405) **(Supplemental Figure 1)**.

**Table 2.** Per-protocol chronic effects of the ActiveBrains physical exercise program on raw and z-score post-exercise (i.e., z-score of change from baseline) sleep-related outcomes.

|  | N<sub>all</sub> | N | Exercise group | N | Mean (95% CI)<br>Control group | Difference between groups<br>(exercise *minus* control) | P |
| --- | --- | --- | --- | --- | --- | --- | --- |
| SRBD scale | 87 | 47 |  | 40 |  |  |  |
| Raw score |  |  | 0.195 (0.165 to 0.225) |  | 0.177 (0.145 to 0.210) | 0.017 (-0.027 to 0.062) | 0.438 |
| z-score |  |  | 0.050 (-0.182 to 0.281) |  | -0.084 (-0.335 to 0.167) | 0.134 (-0.208 to 0.476) |  |
| Total sleep time (min/day) | 66 | 39 |  | 27 |  |  |  |
| Raw score |  |  | 435.231 (422.518 to 447.944) |  | 431.078 (415.799 to 446.357) | 4.152 (-15.725 to 24.029) | 0.678 |
| z-score |  |  | -0.793 (-1.137 to -0.448) |  | -0.905 (-1.319 to -0.491) | 0.113 (-0.426 to 0.651) |  |
| Total time in bed (min/day) | 66 | 39 |  | 27 |  |  |  |
| Raw score |  |  | 504.153 (491.353 to 516.952) |  | 506.982 (491.594 to 522.370) | -2.829 (-22.860 to 17.202) | 0.779 |
| z-score |  |  | -0.799 (-1.202 to -0.395) |  | -0.709 (-1.195 to -0.224) | -0.089 (-0.721 to 0.542) |  |
| Sleep efficiency (%) | 66 | 39 |  | 27 |  |  |  |
| Raw score |  |  | 84.021 (82.744 to 85.298) |  | 82.351 (80.815 to 83.886) | 1.670 (-0.330 to 3.671) | 0.100 |
| z-score |  |  | -0.338 (-0.642 to -0.033) |  | -0.736 (-1.102 to -0.370) | 0.398 (-0.079 to 0.875) |  |
| WASO time (min/day) | 66 | 39 |  | 27 |  |  |  |
| Raw score |  |  | 77.798 (71.222 to 84.374) |  | 88.594 (80.689 to 96.499) | -10.796 (-21.085 to -0.508) | **0.040** |
| z-score |  |  | 0.212 (-0.096 to 0.521) |  | 0.719 (0.348 to 1.090) | -0.507 (-0.989 to -0.024) |  |

z-score values indicate how many standard deviations have the post-exercise program values changed with respect to the baseline mean and standard deviation. E.g., a 0.50 z-score means that the mean value at post-exercise program is 0.50 standard deviations higher than the mean value at baseline, indicating a positive change, with negative values indicating the opposite. All data presented were adjusted for baseline values. SRBD = sleep-related breathing disorders; WASO = wake after sleep onset; SRBD was assessed by the Paediatric Sleep Questionnaire.

### Acute effect of the physical exercise program on sleep

The acute effects on device-assessed sleep outcomes of attending (n=127) vs. not attending (n=73) to the ActiveBrains physical exercise sessions are presented in **Figure 2**. Attending to the physical exercise sessions resulted in a longer sleeping time by 8 min/day (P=0.166), total time in bed by 2 min/day (P=0.750) and better sleep efficiency by 1% (P=0.146); and a reduced -6 min/day WASO time (P=0.185) compared to non-attendance days, although these associations did not reach statistical significance. The SPM analysis of the 24h activity curve shows how attendance days to exercise intervention significantly increased their activity levels compared with the non-attendance days (**Figure 3**).

**Figure 2.**
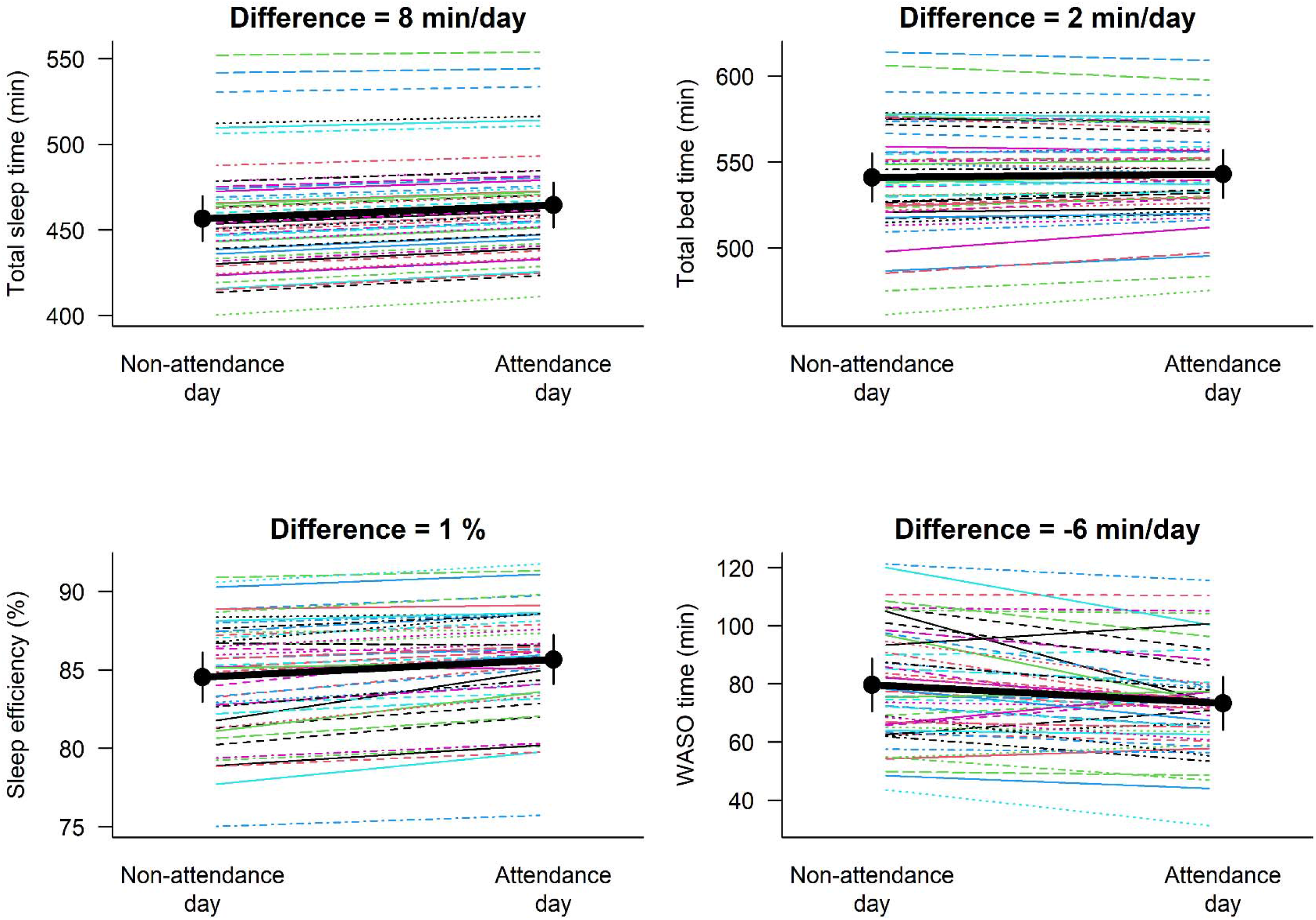
Mean differences for the acute effects four days (excluding Friday, Saturday and Sunday) of the *ActiveBrains* physical exercise program between the attendance days (n=127) vs non-attendance days (n=73) on device-assessed sleep outcomes. WASO = wake after sleep onset. Differences between attendance days vs non-attendance days were non-significant (P>0.05)

**Figure 3.**
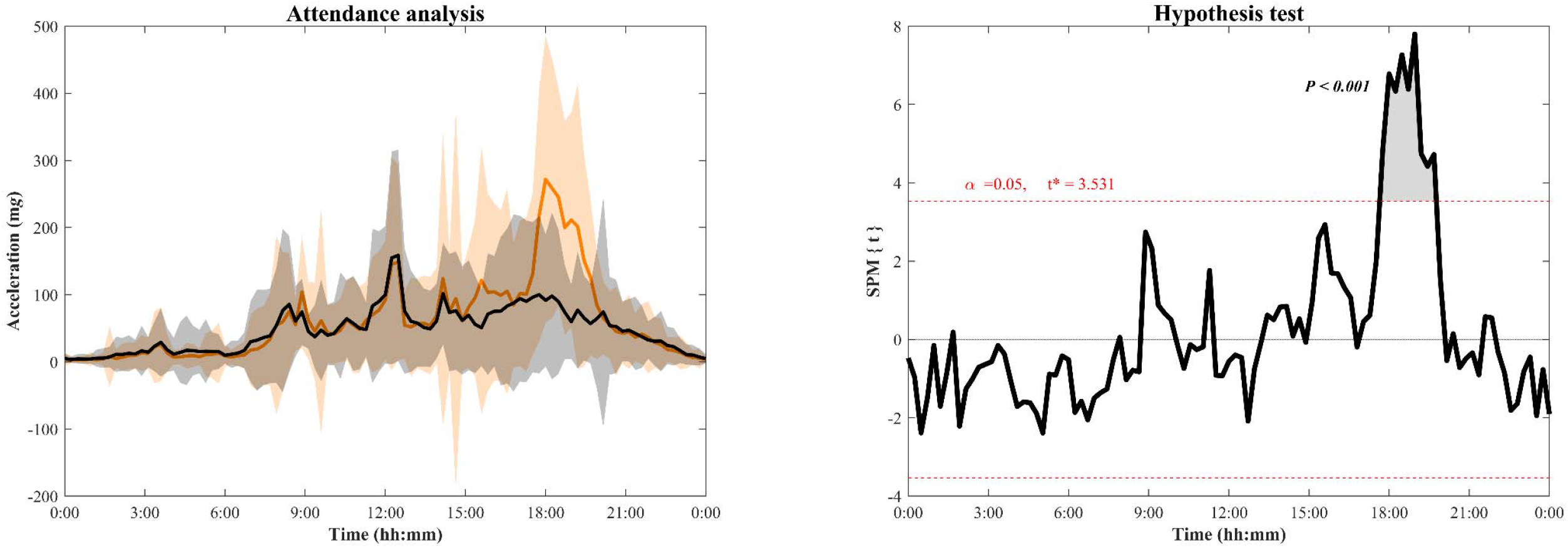
Comparison of means acceleration curves and standard deviation (shaded area) for the attendance (orange line) and non-attendance days (black line) in the exercise group participant during the 10-week of the intervention (excluding Friday, Saturday and Sunday). The hypothesis test shows the threshold (t* = 3.531) at which there are significant physical activity patterns’ differences between attendance and non-attendance exercise days. SPM=Statistical parametric mapping.

## Discussion

### Main findings

The ActiveBrains RCT contributes to the existing literature with the following findings: 1) a 20-week physical exercise program reduces the WASO by around 10 min/day and might increase the sleep efficiency by ∼2% in children with overweight or obesity; 2) all device-assessed sleep indicators, especially the sleep time and the WASO time, were improved in those days in which the participants attended to the sessions, although did not reach statistical significance and it should be taken with caution.

This is one of the few RCT in the literature examining both chronic and acute effects of a physical exercise program on device-assessed sleep habits in childhood obesity. This allows to suggest that exercise has chronic and certain acute benefits (yet non-significant) on sleep habits, specifically on WASO (−6-10 minutes per night) and sleep time (+8 minutes per night). On the other hand, we did not observe a statistically significant effect on SDB.

### Chronic effect of the physical exercise program on sleep

To date, only one RCT investigated the effect of physical intervention on device-assessed sleep in children, but with autism spectrum disorders.^14^ In line with our study, Tse et al. found that a 12-week physical exercise intervention (45 min/session and 2 session/week) had a positive effect on WASO and sleep efficiency.^14^ They also found a significant effect on sleep duration measured by non-dominant wrist-worn accelerometry.^14^ Moreover, in consonance with our results, but using a questionnaire to measure sleep quality, an RCT in children and adolescents reported that a 12-weeks of both judo or ball games benefited sleep quality (i.e., aspects related to sleep pattern and the frequency with which these may occur).^37^ In adolescents with obesity, and in contrast with our findings, a 12-week physical exercise intervention (180 min/week) significantly increased sleep efficiency (0.7 SDs) and total sleep time (0.8 SDs).^15^ In our study, the effect of the intervention was significant only on WASO time, yet certain evidence of effects on sleep efficiency was observed. A null effect was observed on sleep duration in contrast with the significant effects observed in both previous studies.^14,15^ A possible explanation of these inconsistencies could be: 1) this study participants had a healthy sleep at baseline, so there was little room for improvement; 2) children with autism spectrum disorder might have unhealthier sleep habits than children with normal development,^38^ which could enhance the exercise effects; 3) the age groups (adolescents vs. children), and the methods used to measure sleep (polysomnography vs. accelerometry)^15^could lead to inconsistent findings.^39^ Additionally, the type of aerobic exercise (cycling/treadmill/rower vs. running games) or the inclusion or not of resistance exercise might partially explain the differences. A systematic review and meta-analysis in healthy participants concluded that running exercise was related to greater WASO reduction compared to cycling^40^ and, therefore, it could be that inflammation and/or muscular damage caused by this type of aerobic exercise (i.e., running) may positively affect WASO time. However, further investigations are required to elucidate the role of inflammation or muscular damage on sleep.

In regard to sleep disorders, the ActiveBrains exercise program did not improve SDB in children with overweight/obesity. Similarly, in a recent study in obese children with SDB, neither 8-weeks nor 16-weeks of exercise (24 min of HIIT plus 20 min of resistance training) influenced the SRBD scale or subscales.^19^ Consistent with these findings, a previous study in adolescents with obesity observed that apnea-hypopnea index (AHI, measured with polysomnography), an index used to indicate the severity of sleep apnea, was not changed after a 12-week exercise program (180 min/week).^15^ Likewise, Roche et al. found that a combination of 9-months of aerobic exercise and diet had no effect on OSA measured with polysomnography in adolescents with obesity.^41^ However, Davis et al. found that 15 weeks of aerobic exercise (i.e., low-dose exercise of 20 min/day or high-dose of 40 minutes/day) could improve SRBD scale, specifically snoring, in 7- to 11-year-old children with overweight.^18^ The lack of significant effects in our study might be due to the lower SDB severity in our sample at baseline (score of ∼0.19 in a scale range 0 to 1, and 16.2% of our participants presented SDB) compared to in the study by Davis et al. (score of ∼0.24, and 25% of participants presented SDB). Further studies in a sample with higher SDB severity at baseline should confirm or contrast our findings. Furthermore, a current meta-analysis highlights the importance of future well-designed RCTs to understand the independent effects of exercise on OSA and sleep health in children and adolescents with obesity.^42^

### Acute effect of the physical exercise program on sleep

Our results complement the existing literature in normal-weight children ^43^ by suggesting that acute exercise during the afternoon-evening could improve sleep in children with overweight/obesity. It seems that acute aerobic exercise performed at high-intensity have positive effects on sleep efficiency in children.^43^ Our findings are in line with the previous literature by confirming this acute effect (yet non-significant) in children with overweight/obesity enrolled in a concurrent aerobic plus strength exercise program. Likewise, in a study in adolescent girls with obesity, the authors showed that acute exercise in the morning has a positive effect on sleep duration and quality measured with accelerometers.^20^ Furthermore, it seems that acute exercise performed in the evening does not have an adverse effect on sleep, rather a small positive effect in healthy adults.^40^ It could be that children sleep better due to the exhaustion caused by the exercise, independently of the time of the day when it is performed. However, a previous study showed that exercise was beneficial to improve device-assessed sleep (i.e., decreased WASO time and increased sleep efficiency) only when it was completed 4 hours before bed-time and not 2 hours (i.e., late evening) in healthy young adults.^44^ We did not have the opportunity to investigate the timing effect of the exercise on sleep since all participants performed the ActiveBrains exercise sessions at the same time, and we encourage future studies to address that gap in youth. Thus far, the literature in children and youth has examined whether the lifestyle physical activity was associated with sleep the subsequent night’s, finding limited evidence to support this relationship.^17^

### Limitations and strengths

The limitations of this study include: (i) our findings apply to children with overweight/obesity and should not be extrapolated to other populations (e.g., other age groups, normal-weight individuals); (ii) we used questionnaires instead of the gold-standard to measure SDB (polysomnography); (iii) the accelerometer estimates of sleep are based on movement patterns, not purely sleep, yet the accelerometers provide a non-invasive, objective, and valid assessment of sleep in free-living conditions.^31,45^

On the other hand, there are some strengths that should be noted. The randomized design, the use of device-assessed sleep behaviour with accelerometers across an entire week, the advance processing on accelerometer raw data, and the focus on children with overweight/obesity, who often show poorer sleep patterns than normal-weight children. The novel analyses of accelerometer-measured physical activity all throughout the day (24 h) to accurately investigate whether changes in overall physical activity levels took place in the attendance days to physical exercise program and at what time of the day.

## Conclusion

Our findings suggest that exercise can have positive effects on device-assessed sleep habits, particularly chronic effects with certain changes being observed also acutely, by producing ∼6-10 minutes lower WASO time per night. No effects were however observed on SDB in children with overweight/obesity. These findings support public health initiatives that promote physical exercise programs in children with overweight/obesity have the potential to improve their sleep quality. Further RCTs, especially in children with sleep difficulties at baseline are needed to better appreciate the role of exercise in sleep health.

## Supporting information

CONSORT checklist

## Data Availability

We did not obtain children parents consent to widely share the data nor was it included in the IRB protocol.

## Abbreviations

OSA: obstructive sleep apnea,
RCT: randomized clinical trial,
SD: standard deviations,
SDB: sleep-disordered breathing,
SRBD: sleep-related breathing disorders,
WASO: wake after sleep onset.

## Financial disclosure

The authors have indicated they have no financial relationships relevant to this article to disclose.

## Acknowledgments

This work is part of a PhD thesis conducted in the Official Doctoral Programme in Biomedicine of the University of Granada, Spain. The ActiveBrains project was funded by the Spanish Ministry of Economy and Competitiveness and the “Fondo Europeo de Desarrollo Regional (FEDER)” (DEP2013-47540, DEP2016-79512-R, DEP2017-91544-EXP and RYC-2011-09011). L.V.T.-L. is supported by a Grant from the Spanish Ministry of Science, Innovation and Universities (FPU17/04802). C.C.-S. is supported by the Spanish Ministry of Science and Innovation (FJC2018-037925-I). Additional support was obtained from the Alicia Koplowitz Foundation (ALICIAK-2018), the University of Granada, Plan Propio de Investigación 2016, Excellence actions: Units of Excellence, Scientific Excellence Unit on Exercise and Health (UCEES), by the Junta de Andalucía, Consejería de Conocimiento, Investigación y Universidades, and European Regional Development Funds (ref. SOMM17/6107/UGR). In addition, funding was provided by the SAMID III network, RETICS, funded by the PNI + D + I 2017–2021 (Spain), ISCIII-Sub-Directorate General for Research Assessment and Promotion, the European Regional Development Fund (ERDF) (Ref. RD16/0022), the EXERNET Research Network on Exercise and Health (DEP2005-00046/ACTI; 09/UPB/19; 45/UPB/20; 27/UPB/21), the European Union’s 2020 research and innovation program under grant agreement No.667302, and the HL-PIVOT network-Healthy Living for Pandemic Event Protection. Additional funding was obtained from the Andalusian Operational Programme supported with European Regional Development Funds (ERDF in English, FEDER in Spanish, project ref: B-CTS-355-UGR18).

## Contributors’ Statement

Ms Torres-Lopez drafted the initial manuscript, conducted all statistical analyses, reviewed and revised the manuscript. Drs. Migueles and Cadenas-Sanchez collected data, carried out the initial analyses and reviewed and revised the manuscript. Dr. Bendtsen participated in analysis and critically reviewed and revised the manuscript. Drs. Henriksson, Mora-Gonzalez, Löf and Chaput critically reviewed the manuscript for important intellectual content. Dr. Ortega was the principal investigator of this study and conceptualized and designed the study, coordinated and supervised data collection and critically reviewed and revised the manuscript. All authors approved the final manuscript as submitted and agree to be accountable for all aspects of the work.

## Notes

### Competing Interest Statement

The authors have declared no competing interest.

### Clinical Trial

NCT02295072

### Author Declarations

The ActiveBrains RCT was approved by the Human Research Ethics Committee of the University of Granada.

