## Supplementary material for "Effects of exercise on sleep in children with overweight/obesity: A randomized clinical trial": CONSORT checklist

### Checklist of items to include when reporting a randomized trial (56-58)

| PAPER SECTION And topic | Item | Description | Reported on page # |
|---|---|---|---|
| *TITLE & ABSTRACT* | 1 | How participants were allocated to interventions (*e.g.*, "random allocation", "randomized", or "randomly assigned"). | 1 and 3 |
| *INTRODUCTION* Background | 2 | Scientific background and explanation of rationale. | 4-5 |
| *METHODS* Participants | 3 | Eligibility criteria for participants and the settings and locations where the data were collected. | 5-6 |
| Interventions | 4 | Precise details of the interventions intended for each group and how and when they were actually administered. | 5-6 |
| Objectives | 5 | Specific objectives and hypotheses. | 5 |
| Outcomes | 6 | Clearly defined primary and secondary outcome measures and, when applicable, any methods used to enhance the quality of measurements (*e.g.*, multiple observations, training of assessors). | 6-7 |
| Sample size | 7 | How sample size was determined and, when applicable, explanation of any interim analyses and stopping rules. | 5 |
| Randomization -- Sequence generation | 8 | Method used to generate the random allocation sequence, including details of any restriction (*e.g.*, blocking, stratification). | 5 |
| Randomization -- Allocation concealment | 9 | Method used to implement the random allocation sequence (*e.g.*, numbered containers or central telephone), clarifying whether the sequence was concealed until interventions were assigned. | - |
| Randomization -- Implementation | 10 | Who generated the allocation sequence, who enrolled participants, and who assigned participants to their groups. | 5 |
| Blinding (masking) | 11 | Whether or not participants, those administering the interventions, and those assessing the outcomes were blinded to group assignment. When relevant, how the success of blinding was evaluated. | 6 |
| Statistical methods | 12 | Statistical methods used to compare groups for primary outcome(s); Methods for additional analyses, such as subgroup analyses and adjusted analyses. | 7-9 |
| RESULTS Participant flow | 13 | Flow of participants through each stage (a diagram is strongly recommended). Specifically, for each group report the numbers of participants randomly assigned, receiving intended treatment, completing the study protocol, and analyzed for the primary outcome. Describe protocol deviations from study as planned, together with reasons. | Figure 1 |
| Recruitment | 14 | Dates defining the periods of recruitment and follow-up. | 5 |
| Baseline data | 15 | Baseline demographic and clinical characteristics of each group. | Table 1 |
| Numbers analyzed | 16 | Number of participants (denominator) in each group included in each analysis and whether the analysis was by "intention-to-treat" . State the results in absolute numbers when feasible (*e.g.*, 10/20, not 50%). | 9 and 10 |
| Outcomes and estimation | 17 | For each primary and secondary outcome, a summary of results for each group, and the estimated effect size and its precision (*e.g.*, 95% confidence interval). | 10 |
| Ancillary analyses | 18 | Address multiplicity by reporting any other analyses performed, including subgroup analyses and adjusted analyses, indicating those pre-specified and those exploratory. | 10 |
| Adverse events | 19 | All important adverse events or side effects in each intervention group. | - |
| DISCUSSION Interpretation | 20 | Interpretation of the results, taking into account study hypotheses, sources of potential bias or imprecision and the dangers associated with multiplicity of analyses and outcomes. | 11-14 |
| Generalizability | 21 | Generalizability (external validity) of the trial findings. | 11-14 |
| Overall evidence | 22 | General interpretation of the results in the context of current evidence. | 11-14 |
