## Supplemental Table 1 and Supplemental Figure 1 for "Effects of exercise on sleep in children with overweight/obesity: A randomized clinical trial"

**Table S1.** Intention-to-treat chronic effects of the ActiveBrains physical exercise program on raw and z-score post-exercise (i.e., z-score of change from baseline) sleep-disorders outcomes.

| | | | Mean (95% CI) | | | | |
|---|---|---|---|---|---|---|---|
| | $N_{all}$ | N | Intervention group | N | Control group | Difference between groups (Intervention *minus* control) | *P* |
| SRBD scale | | | | | | | |
| Raw score | 109 | 57 | 0.193 (0.167 to 0.220) | 52 | 0.189 (0.162 to 0.217) | 0.004 (-0.034 to 0.042) | 0.844 |
| z-score | | | 0.045 (-0.158 to 0.247) | | 0.016 (-0.196 to 0.227) | 0.029 (-0.264 to 0.322) | |
| Total sleep time | 109 | 57 | | 52 | | | |
| Raw score | | | 437.516 (429.021 to 446.011) | | 431.294 (422.400 to 440.189) | 6.221 (-6.087 to 18.529) | 0.319 |
| z-score | | | -0.665 (-0.906 to -0.424) | | -0.841 (-1.094 to -0.589) | 0.176 (-0.173 to 0.525) | |
| Total time in bed | 109 | 57 | | 52 | | | |
| Raw score | | | 501.652 (491.910 to 511.394) | | 506.378 (496.176 to 516.580) | -4.726 (-18.874 to 9.422) | 0.509 |
| z-score | | | -0.888 (-1.198 to -0.578) | | -0.738 (-1.063 to -0.413) | -0.150 (-0.601 to 0.300) | |
| Sleep efficiency | 109 | 57 | | 52 | | | |
| Raw score | | | 83.906 (82.913 to 84.899) | | 82.843 (81.803 to 83.882) | 1.063 (-0.374 to 2.501) | 0.145 |
| z-score | | | -0.167 (-0.375 to 0.040) | | -0.390 (-0.607 to -0.172) | 0.222 (-0.078 to 0.523) | |
| WASO time | 109 | 57 | | 52 | | | |
| Raw score | | | 79.681 (74.502 to 84.860) | | 84.817 (79.395 to 90.239) | -5.136 (-12.635 to 2.363) | 0.177 |
| z-score | | | 0.118 (-0.104 to 0.341) | | 0.339 (0.106 to 0.572) | -0.221 (-0.543 to 0.102) | |

z-score values indicate how many standard deviations have the post-exercise program values changed with respect to the baseline mean and standard deviation. E.g., a 0.50 z-score means that the mean value at post-exercise program is 0.50 standard deviations higher than the mean value at baseline, indicating a positive change, with negative values indicating the opposite. All data presented were adjusted for baseline values. SRBD = sleep-related breathing disorders; WASO = wake after sleep onset.

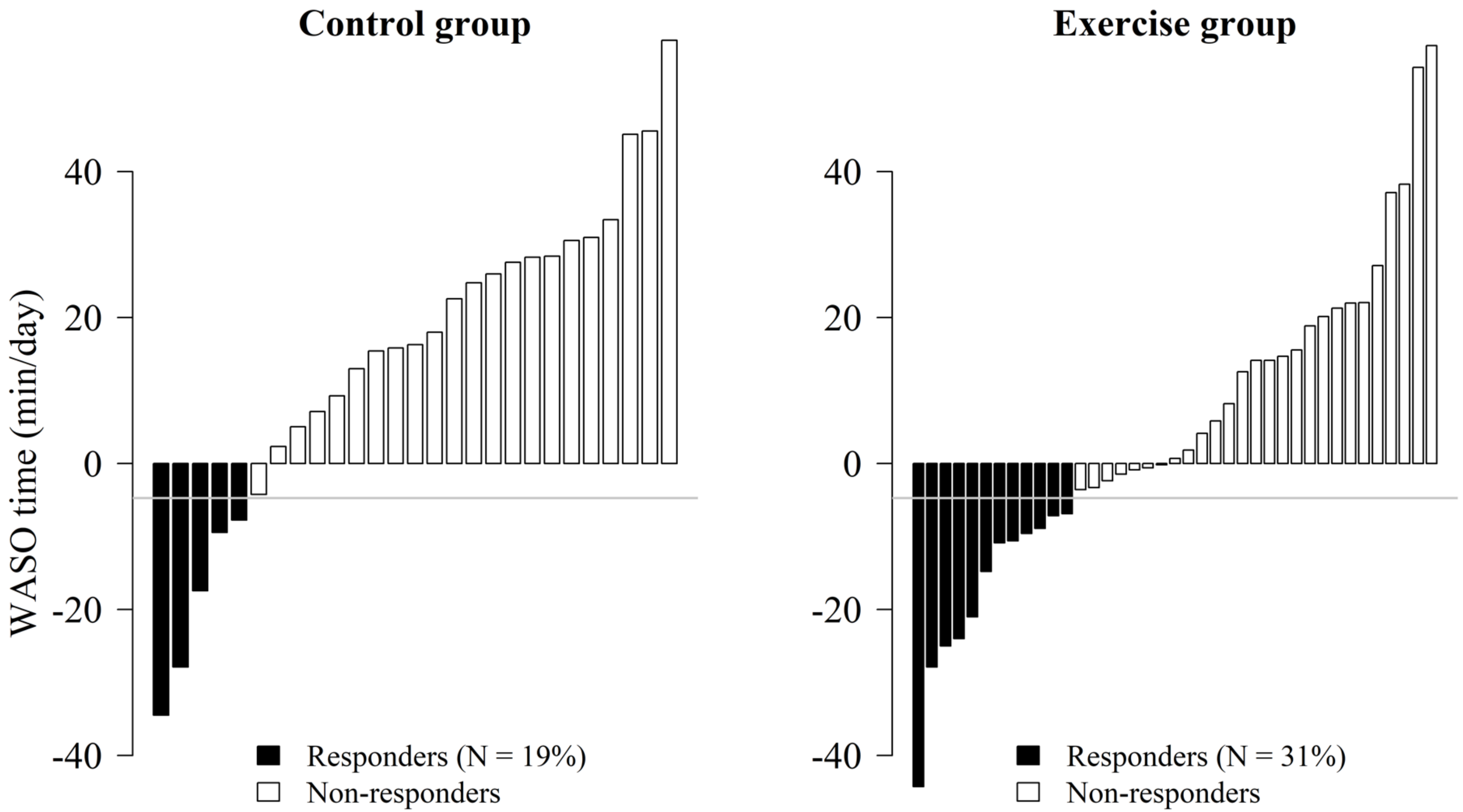


**Figure S1.** Pre-post change distribution in the outcome significantly affected by the exercise program (i.e., wake after sleep onset time, WASO time). Data analyses were primarily conducted under the per-protocol principle, i.e., attending to 70% of the sessions. Participants were classified as responders when they reduce the WASO time after the intervention (Cohen's d ≥ 0.2), whilst non-responders were categorized for those participants who did not experience a reduction (Cohen's d <0.2).
